# The NPIS Model: A Standardized, Consensus-Based Framework for Evaluating Non-Pharmacological Interventions

**DOI:** 10.1101/2025.04.04.25325250

**Authors:** Gregory Ninot, Emeline Descamps, Ghislaine Achalid, Sébastien Abad, Fabrice Berna, Christine Belhomme, François Carbonnel, Patrizia Carrieri, Patricia Dargent-Molina, Frederic Fiteni, Aude-Marie Foucaut, Alice Guyon, Arnaud Legout, Beatrice Lognos, Nicolas Molinari, Julien Nizard, Michel Nogues, François Paille, Pierrick Poisbeau, Lise Rochaix, Bruno Falissard

## Abstract

**Background:** The term non-pharmacological intervention (NPI) refers to health prevention and care protocols supervised by healthcare professionals. However, no precise and widely adopted definition currently exists. Additionally, unlike drugs, NPIs lack a global evaluation framework due to the absence of consensus among stakeholders, driven by the heterogeneity of intervention content and study protocols. This heterogeneity limits scientific impact, hinders dissemination and continuous improvement of practices, and contributes to significant mistrust among professionals and users.

**Method:** From 2022 to 2023, we conducted a consensus study, collaborating with over 1,000 stakeholders—including researchers, healthcare users, healthcare practitioners, health operators, scientific societies and health authorities—to co-construct a NPI definition along with a framework for evaluating NPIs that aligns with their specific characteristics and international health research standards. This framework—the NPIS Model—was developed under the guidance of a multidisciplinary committee of 22 experts through iterative, open, and documented exchanges across four stages: i) creation of an initial list of ethical and methodological items by a committee of 70 members, ii) refinement of this list by a committee of 300 members, iii) open voting on each item by 503 voters, and iv) consultation with 36 scientific societies and 14 health authorities.

**Findings:** We reached a consensus defining an NPI as an *“evidence-based, effective, personalized, non-invasive health prevention or care protocol, registered and supervised by a qualified professional”*. Based on this definition, we developed the NPIS Model, which includes 77 recommendations for evaluating NPIs—14 ethical and 63 methodological. Each recommendation received at least 80% agreement among voters. The recommendation are categorized into five types of studies: mechanistic, observational, prototypical, intervention, and implementation. To date, the NPIS Model has been endorsed by 31 scientific societies and three health authorities.

**Interpretation:** The NPIS Model promotes transparency, methodological rigor, ethical standards, and transferability for NPI research, increasing the value of NPIs for researchers, healthcare practitioners, healthcare users, and health authorities.

**Funding:** National Institute of Health and Medical Research (INSERM) “*Fonds d’amorçage recherche participative”*.

**Research in context:** *Evidence before this study:* Publications on non-pharmacological interventions (NPIs) are rapidly increasing. We performed a MEDLINE (Pubmed) search on June 16, 2023, using the following request on all fields: “non-pharmacologic” OR “nonpharmacologic” OR “non-pharmacological” OR “nonpharmacological” OR “non-pharmaceutical” OR “nonpharmaceutical” OR “non-medication” OR “non-drug” OR “nondrug”. This search identified 33,387 articles, with a fourfold increase in publications over the past decade. Despite this growth, no consensus-based framework exists for conducting NPI research. Alongside our PubMed search, we reviewed all guidelines in the EQUATOR network. Although some guidelines cover specific aspects of NPI studies, we found no consensus-based evaluation framework tailored specifically to NPIs.

*Added value of this study:* We co-constructed an NPI definition along with a framework—the NPIS Model—in a consensus study involving over 1,000 stakeholders, including researchers, healthcare users, healthcare practitioners, health operators, scientific societies and health authorities. Extensive stakeholder engagement has shaped the NPIS Model to reflect a broad range of perspectives, ensuring its relevance and acceptance. The NPIS Model contains 14 ethical items and 63 methodological items, each achieving over 80% agreement. It has received support from 31 scientific societies and three health authorities, which underlines its relevance across health sectors.

*Implications of all the available evidence:* A consensus-based evaluation framework for NPIs will significantly enhance the quality of NPI research by improving consistency, reproducibility, and comparability. The NPIS Model promotes methodological rigor, ethical standards, and research transferability. Ultimately, we expect this model to benefit researchers, healthcare practitioners and users, and inform public health policies.

## Introduction

Over the past 20 years, non-pharmacological interventions (NPIs) for human health have gained interest not only among the general public but also among healthcare practitioners and health authorities.^2^ These patient-centered approaches are increasingly implemented in both prevention and care across various healthcare settings, including chronic diseases management, rehabilitation medicine, mental health, childcare, addiction treatment, occupational medicine, disability support, gerontology, and end-of-life care.^2^ Since 2003, even the World Health Organization (WHO) has contributed to the development and promotion of NPIs.^16–18^

### The challenge of defining NPIs

Despite this growing recognition, there is still no universally accepted definition of NPIs.^46^ Different publications include or exclude various types of interventions, leading to inconsistencies in scope. For instance, some sources classify surgery or medical devices as NPIs,^4^ while others explicitly exclude them.^3^ This variability in definitions complicates the evaluation process, as assessing a surgical procedure involves different methodological challenges than assessing a medical device or a physical therapy. To address this issue, in 2011, the Haute Autorité de Santé (HAS)—France’s highest health authority for evaluating and recommending health practices and technologies—published a report recommending that the term NPI be used for interventions related to hygiene and dietary rules, psychological treatments, and physical therapies.^45^ Building on this authoritative recommendation, we adopted the term NPI and further refined its definition through a consensus-based process, aligning with the latest recommendations in the litterature.^46^ The final consensus, as described in this work, defines an NPI as: “an evidence-based, effective, personalized, non-invasive health prevention or care protocol, registered and supervised by a qualified professional”.

### The need for a comprehensive evaluation framework

Beyond defining NPIs, a critical challenge remains: establishing a structured, consensus-based framework to guide their scientific evaluation. Unlike drugs, which follow a well-established evaluation pipeline from preclinical research to regulatory approval,^23–29^ NPIs require a broader and more integrative assessment approach. Their evaluation must account for not only efficacy but also factors such as standardization of the intervention, feasibility, ethical considerations, mechanisms of action, and long-term impact on health outcomes.

Efforts are underway to establish an evaluation framework for NPIs,^33,39,40^ however, these initiatives are in their early stages and remain focused on a single aspect of NPI evaluation. International recommendations, such as those in the EQUATOR Network, provide guidance on specific aspects of NPI evaluation, including the CONSORT statement on how to design a randomized control trial for NPIs^4,7^ or, recently, the CoPPS statement on how to create a control group for clinical or mechanistic studies for physical, psychological, and self-management therapies.^3^ However, these guidelines focus primarily on randomized controlled trials (RCTs) and do not constitute a global evaluation framework encompassing all essential study types required for a robust assessment of NPIs. Furthermore, existing frameworks such as Orbit^6^ and CONSORT^7^ models are inspired by drug evaluation, prioritizing internal validity (i.e., absolute comparability of groups) over external validity (i.e., capacity to validate the results found for the target population) that is crucial for NPIs. Other frameworks are based on theories of behavioral change,^8^ are inspired by engineering,^9^ or propose hybridizations such as Most.^10^ Few consider a patient-centered approach or the broader context in which NPIs are applied.

A 2019 systematic review identified 46 different frameworks for evaluating behavioral intervention technologies,^5^ yet none has achieved widespread adoption or consensus. The lack of a unified framework contributes to fragmented evaluation approaches, limiting comparability across studies, hindering reproducibility, and reducing the transferability of findings into clinical practice.^11-13^ As a result, scientific rigor in NPI research remains inconsistent, which in turn weakens the credibility of NPIs in healthcare decision-making.^2^ This gap also opens the door to the proliferation of pseudoscientific alternative practices^14^ and the intensification of the tension between evidence-based medicine and approaches based on personal experience and tradition.^15^

### A consensus-driven approach to NPI evaluation

Both researchers^5,7,11,19–21^ and health authorities^13,16,22^ have called for the establishment of a comprehensive evaluation framework that systematically integrates all aspects of NPI assessment. To be effective, such a framework must be: i) consensus-based, ensuring alignment among all stakeholders, including researchers, healthcare users, healthcare practitioners, health operators, scientific societies, and health authorities; ii) comprehensive, covering all study types, from defining the intervention to evaluating its real-world implementation; iii) adapted to NPIs, recognizing their specific characteristics while adhering to established health research standards.

This study aims to develop the first consensus-driven framework for the scientific evaluation of NPIs, engaging all stakeholders to ensure alignment with international ethical and methodological research standards while addressing the unique characteristics of NPIs.

## Methods

### General principle

We conducted a preliminary epistemological work on an NPI evaluation framework from 2011 to 2020 as part of a university collaborative platform in Montpellier, France.^2^ We extended this work with the goal of establishing a pragmatic and global model for evaluating NPIs based on the principles of evidence-based medicine and patient-centered approach. We structured this work around five types of studies (see Figure 1): (i) mechanistic studies that highlight the biological mechanisms and active psychosocial processes of an NPI, (ii) observational studies that monitor the evolution of practices related to an NPI (iii) prototypical studies that describe all the characteristics of an NPI, (iv) intervention studies, also called clinical studies, that determine the effectiveness of an NPI on health markers of a target population, and finally, (v) implementation studies that verify the conditions for deploying an NPI in a specific setting, and related modalities to adapt the intervention to the context.

**Figure 1:**
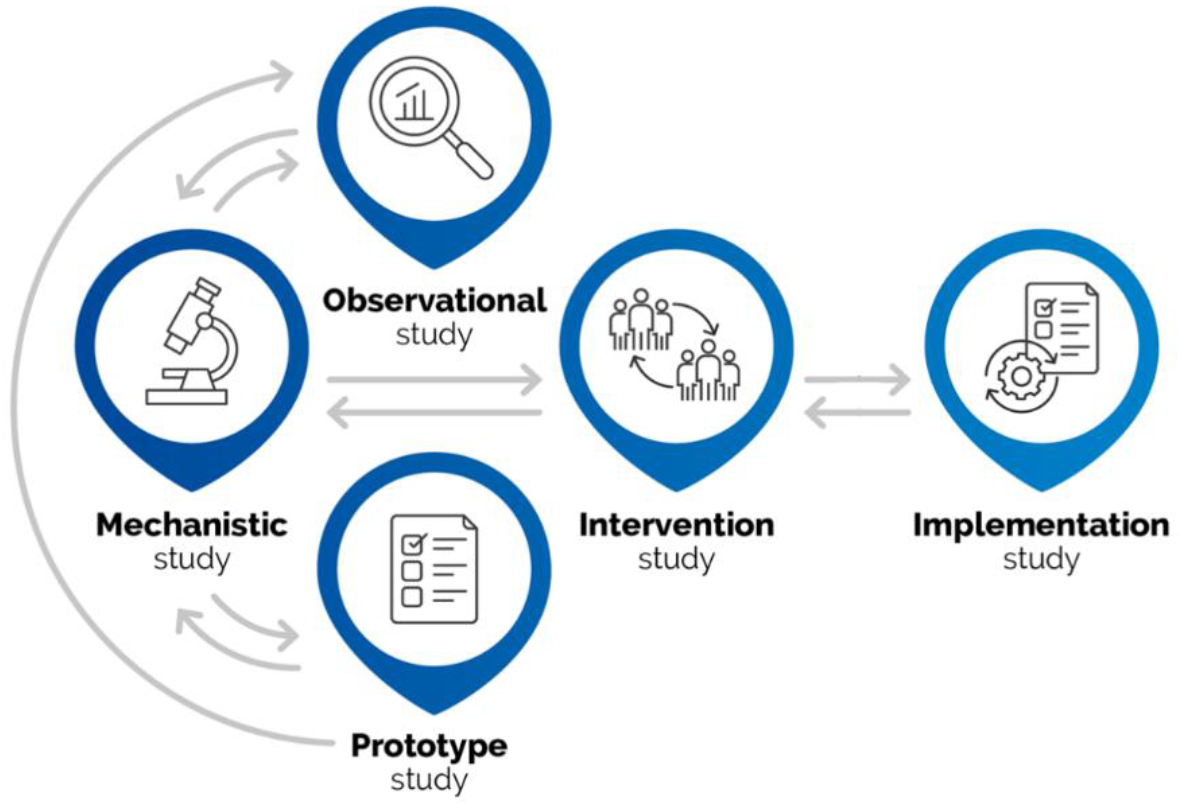
The five types of studies covered by NPIS Model and their dependencies.

Following this preliminary work, the NPI Society was created in 2021 in Montpellier, France. It is a non-profit scientific society dedicated to fostering research and development on NPI, with its first mission being the development of the NPIS Model. Since its creation, it has been open to international members, including researchers, healthcare practitioners, health operators, and healthcare user representatives from other European countries, as well as North America.

The NPIS Model was developed under the supervision of a steering committee composed of Ghislaine Achalid (healthcare user representative), Emeline Descamps (researcher), and Gregory Ninot (professor). The steering committee issued an open call to all NPI Society members to create an expert committee. The inclusion criterion required recognized expertise in a field relevant to the creation of the NPIS Model. The steering committee assessed the relevance and level of expertise. The exclusion criterion was any conflict of interest with a pharmaceutical industry. The expert committee consisted of 22 members, including 20 scientific experts and 2 healthcare user representatives (see Annex D for the complete list with affiliation and domain of competencies). During the development, we followed the EQUATOR recommendations^23^ and the Appraisal of Guidelines, Research and Evaluation (AGREE II) checklist^24^ (see Annex F for completed checklists). In particular, to select and adapt items, we involved all types of academic and non-academic stakeholders: researchers, healthcare practitioners, healthcare users, and health operators. We informed scientific societies and authorities before defining the NPIS Model and we consulted them after the definition for their opinions. In summary, we co-constructed a model consisting of recommendation items with iterative and open exchanges with all stakeholders.

The entire study was conducted remotely using videoconferencing and online voting, due to the on-going impact of the COVID-19 pandemic, which significantly restricted professional travel. Despite this, we observed no adverse impact on the study.

In the following, we detail the four successive steps taken between 2022 and 2023 to construct the NPIS Model. Figure 2 provides an overview of these steps.

**Figure 2:**
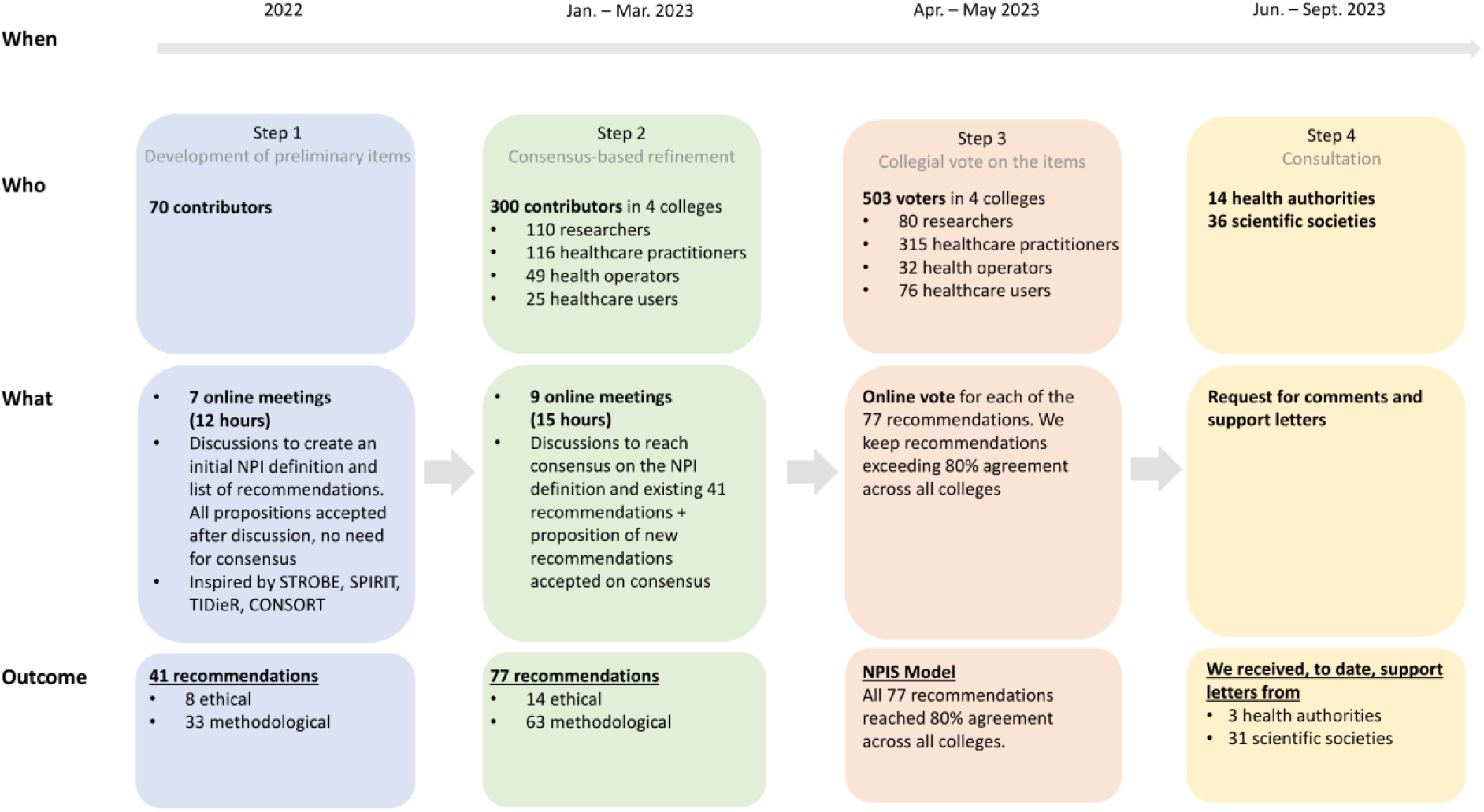
Overview of the methodology and results.

### Step 1: Development of preliminary recommendation items

In 2022, we developed a preliminary definition for NPIs, along with methodological and ethical recommendation items. To recruit contributors for this task, the steering committee issued an open call via the NPI Society^30^ newsletter and emails to all NPI Society members. The inclusion criteria were being member of the NPI Society and francophone to facilitate the communication among contributors. There were no exclusion criteria or no limit on the number of contributors.

We organized seven interdisciplinary and intersectoral consultations through online meetings, totaling 12 hours, with all contributors attending for the full duration. The goal was both to establish a clear definition for NPI and to develop a list of recommendations for the five study types (see Figure 1), inspired by the literature and adapted to NPIs. These meetings led to the identification of relevant scientific articles^3-5,25-29^ and the development of a shared working document, edited by the expert committee. To be eligible, scientific articles needed to be published in a peer-reviewed journal indexed in a recognized database such as PubMed or Scopus and adhere to the ICMJE recommendations.^41^ During these meetings, all propositions were discussed, but no consensus was sought at this step. The expert committee crafted the definition of NPI based on these discussions, and any member of the expert committee could decide to add a discussed recommendation to the document that became the initial version of the NPIS Model. The recommendation items were inspired by several recommendation standards: i) Strengthening the Reporting of Observational Studies in Epidemiology (STROBE) guidelines,^25^ Standard Protocol Items: Recommendations for Interventional Trials (SPIRIT) 2013 document and its 2022 extension,^26,27^ iii) Template for Intervention Description and Replication (TIDieR) checklist and guide,^28^ iv) Consolidated Standards of Reporting Trials (CONSORT) statement on evidence-based reporting for trials assessing non-pharmacologic treatments,^4^ and v) Standards for Reporting Implementation Studies (STaRI) statement and checklist for implementation studies.^29^ The presentation of the recommendation items followed the PICO criteria: *Population, Intervention, Comparison, and Outcomes*. Finally, we created a website dedicated to the project.^30^

### Step 2: Consensus-based refinement of the recommendation items

Between January and March 2023, we refined the preliminary NPI definition, and the 41 ethical and methodological recommendation items from step 1 through consensus meetings. To recruit contributors for this task, the steering committee issued an open call via emails and social media posts. The inclusion criterion was being francophone and belonging to one of the four colleges, ensuring representation from all stakeholders involved in NPIs: researchers, healthcare practitioners, healthcare users, or health operators. Hereafter, we use “college” exclusively to refer to these four groups. The requirement for membership in the NPI Society was removed, and there were no exclusion criteria or limit on the number of contributors.

We scheduled nine interdisciplinary and intersectoral consultation meetings, covering all study types (see Figure 1), totaling 15 hours. Not all participants attended all 15 hours, but, before each meeting, we shared with all contributors the latest version of the working document on the NPI definition and the ethical and methodological items. During the meetings, the NPI definition and each item was read aloud and revised based on participant consensus. Participants could propose modifications to the definition and suggest new recommendation items, either during a meeting or within the shared working document. To assess consensus, we asked whether anyone objected to the NPI definition or an item, with opposition expressed verbally. If no objections were raised, the definition or the item was considered agreed upon. If opposition arose, those who disagreed were invited to propose modifications, which were then discussed either during the same meeting or scheduled for the next one. Ultimately, consensus was reached for the NPI definition and all discussed items. The steering committee (GN, ED, GA) was responsible for editing the document. Between any two meetings, participants could make suggestions in revision mode in the working document. The expert committee verified the comprehensibility of the NPI definition and all recommended methodological and ethical items. To get additional feedback, we presented the last version of the working document to the NPI Society international congress in Montpellier, on March 22, 2023. During a final meeting, the expert committee validated each ethical and methodological recommendation item and produced the final version of the document to be put to a collegial vote.

### Step 3: Collegial vote on the recommendation items

Between April and May 2023, we organized an online vote on all recommendation items that had been validated by consensus in step 2. We used an online voting platform hosted in France that complies with the GDPR. To recruit voters, the steering committee issued an open call via emails and social media posts. The inclusion criterion was residing in France and being a member of one of the four colleges, with no exclusion criteria or limit on the number of voters. To ensure participants could vote only once, we manually validated each voter’s first name, last name, profession, organization, and email address. Each item had three possible responses: *agree, disagree, no answer*. Voters who responded “*no answer”* were considered not having voted for the relevant item. This choice was justified because, depending on the college, certain items were not relevant to the voter. Therefore, the voter’s choice could be interpreted as a lack of opinion or competence, rather than opposition. We consider a recommendation item to have been validated by vote if there is at least 80% agreement within each college for each item, that is: *agree / (agree + disagree) * 100* ≥ *80*.

### Step 4: Consultation of health authorities and scientific societies

Between June and September 2023, we gathered feedback on each ethical and methodological item from health authorities and 36 scientific societies in the health sector. Since no official list of health authorities and scientific societies was available, the steering committee compiled a list of all known francophone health authorities and scientific societies, as provided by the expert committee. During two meetings on August 28 and September 18, 2023, the expert committee discussed each comment and integrated the relevant ones into the final NPIS Model, which is available in Annex A.

### Impact of item order on voter participation

To assess whether the position of an item in the questionnaire influenced the number of voters, we formulated the null hypothesis: the position of an item has no impact on the number of voters for that item. To test this hypothesis, we conducted a permutation test by randomly shuffling the item order multiple times (n=100,000).^36^ We computed two test statistics: the difference in the total number of “no answer” votes between the first and last item (T1), the difference in the total number of “no answer” votes between the first and last ten items (T2). We calculated two-tailed p-values for both statistics to determine whether any observed differences were significant.

## Results

In Step 1, 70 contributors (including the expert committee) collaborated to develop the initial NPI definition, along with 41 recommendations—8 ethical and 33 methodological—which together formed version 1 of the NPIS Model. Among the contributors, two were from Belgium, two from Canada, and one from Spain.

Step 2 involved 300 contributors, organized into four colleges: 110 researchers, 116 healthcare practitioners, 49 health operators, and 25 healthcare users. This step still included the five non-French contributors. Through nine scheduled meetings, these contributors developed version 2 of the NPIS Model, leading to the final consensus-based definition of NPI and 77 recommendations: 14 ethical and 63 methodological. This increase from 41 to 77 recommendations resulted from refining and splitting the initial recommendations into more specific ones. Another key change concerned the ethical items: initially, there were categorized by study type, but a consensus emerged that ethical considerations apply to all study types and should be defined separately. Additionally, contributors proposed several new ethical items during discussions.

At the end of step 2, we have crafted a consensus-based definition for NPI that is restricted to practices that are primarily bodily (e.g., manual therapies), psychosocial (e.g., psychotherapies), and nutritional (e.g., targeted diets) practices. Specifically, we defined an NPI as an *“evidence-based, effective, personalized, non-invasive health prevention or care protocol, registered and supervised by a qualified professional”*. We discuss the NPI definition in more detail in the Discussion section.

Step 3 involved 503 voters from the four colleges: 80 researchers, 315 healthcare practitioners, 32 health operators, and 76 healthcare users. We provide the breakdown by profession in Annex E. Figure 3 presents the percentages of agreement for each of the 77 recommendations by college. Since all items exceeded 80% agreement across all colleges, they were all included in the NPIS Model.

**Figure 3:**
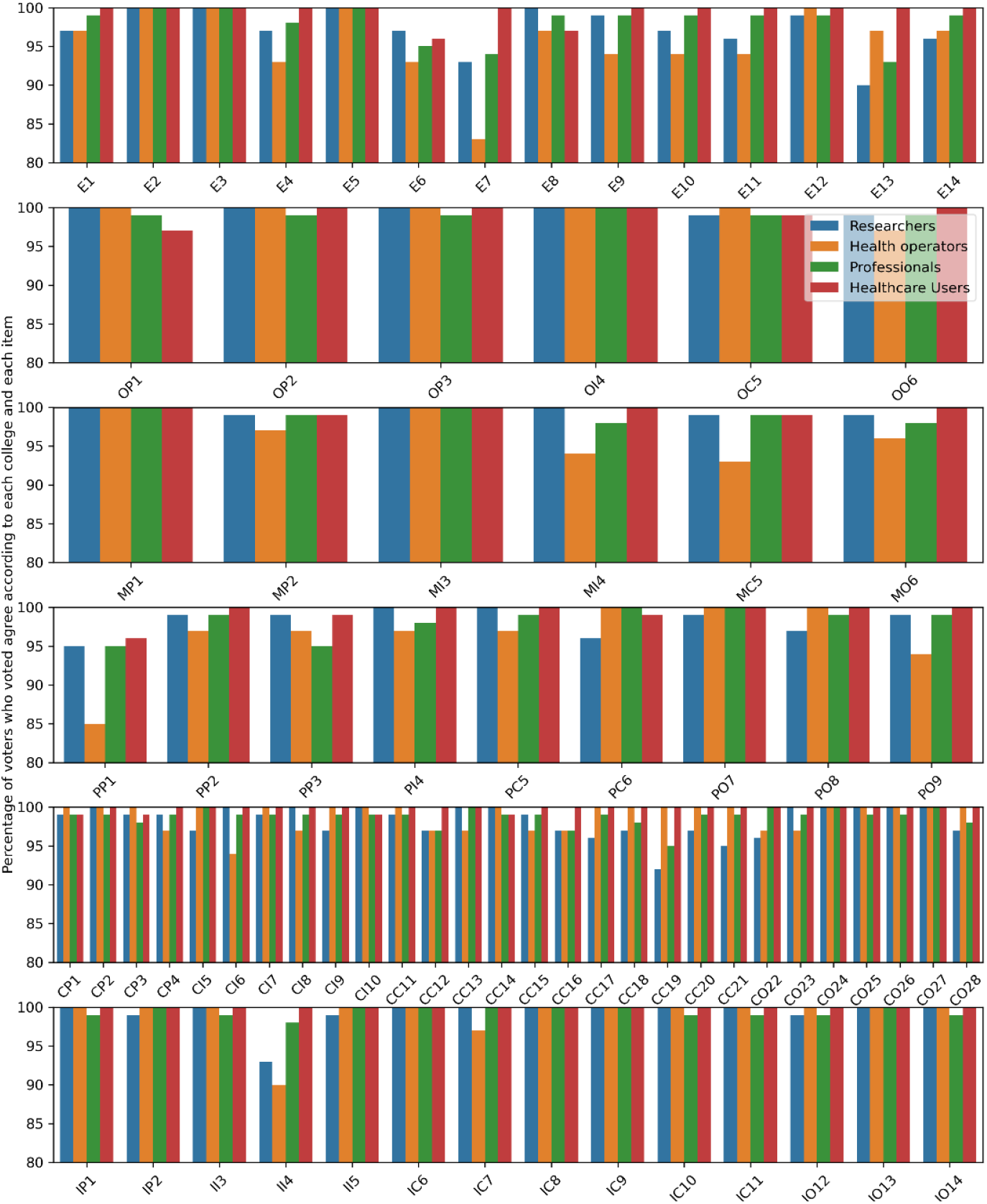
Percentage of voters (n=503) who voted “agree” by college and item. The x-axis represents ethical and methodological items defined in Annex A. Ethical items are labeled “E”, while methodological items use a two-letter code. In this code, the first letter represents the study type: O (Observational), M (Mechanistic), P (Prototypical), C (interventional/Clinical), and I (Implementation). The second letter corresponds to the PICO criteria: P (Population), I (Intervention), C(Comparison), and O (Outcomes). For example, PO8 refers to the eighth Prototypical recommendation item related to an outcome. The top figure represents ethical items, followed by figures for observational study items, mechanistic study items, prototypical study items, intervention study items, and, lastly, implementation study items. The y-axis starts at the 80% threshold.

We found no significant impact of item position on voter participation. The total number of “no answer” votes for the first item was 22, while the last item received 17 votes (T1 = 5). For the first ten items, the total number of “no answer” votes were 331, compared to 257 for the last ten items (T2 = 74). The permutation test confirmed that these differences were not statistically significant (T1 p = 0.81, T2 p = 0.56). Therefore, we failed to reject the null hypothesis, indicating that item order did not meaningfully influence voter response rates.

Voters responded to the large majority of the 77 items: 34% responded to all items, 76% responded to at least 70 items, and 95% responded to at least 60 items. The lowest response rate was 69%. The number of voters who voted to *agree* or *disagree* met the minimum threshold value for all colleges for each item, with agreement surpassing 80% for all items. Consequently, version 3 of the NPIS Model contained 77 recommendations (14 ethical and 63 methodological).

In Step 4, 30 out of 36 scientific societies in the field of health and three out of 14 health authorities participated.

One scientific society for cancer redirected the demand for support and feedback to the French Society of Supportive Care in Oncology, which also endorsed the model. The Geriatrics Society and the Society of Physical and Rehabilitation Medicine redirected their request for support and feedback to their respective European Scientific Societies. The French Society of Thermal Medicine and the French Association of Psychiatry have not responded. The National College of General Practitioner Teachers made a request to reword an item, which was accepted by the expert committee during its first meeting. Specifically, the phrase “and if possible, with a minimal clinically important difference (MCID)” was added to item CO22: “Use objective and subjective criteria (e.g., patient-reported outcomes) employing the SMART approach (Specific, Measurable, Achievable, Realistic, and Timely), measured with validated instruments in the local language”.

Three authorities supported the NPIS Model: the French National Cancer Institute, the National Centre for Palliative and End-of-Life Care, and the French Clinical Research Infrastructure Network. The French Agency for the Sanitary Safety of Food, the Environment, and Work replied that the subject of NPIs was outside its area of expertise. The other French authorities responded that they were not authorized to issue a support letter to a scientific society (Ministry of Health, National Authority for Health, Health Innovation Agency, Miviludes, French Public Health Agency, French Conference of Ethic Committees, Federation of Research Ethic Committees, Health Insurance). The French Academy of Medicine and the High Committee of Public Health have not responded. Following a question from the French Conference of Ethic Committees, the second expert committee meeting focused on the sustainability of the data. However, no modification was made to any of the recommendations. Ultimately, the final version of the online, freely accessible NPIS Model includes 77 recommendations: 14 ethical and 63 methodological. We present the list of recommendations in Table 1. However, they are not meant to be applied as a monolithic block. For instance, a researcher conducting a prototypical study should apply only the 14 ethical items and the 9 items specific to prototypical studies (respectively green and blue items in Table 1). Researchers and clinicians should refer to the Discussion section for guidance on applying the NPIS Model.

**Table 1:** Final ethical and methodological recommendation items for the NPIS Model. These items are categorized into 6 sections: ethical items in green, observational study items in orange, mechanistic study items in pink, prototypical study items in blue, intervention study items in yellow, and implementation study items in gray. The detailed explanation of the code column is given in the caption of Figure 3. A detailed presentation of the 77 items with explanations is available in Annex A.

| Code | Item |
| --- | --- |
| E1 | Respect the laws, regulations and ethics charters of the research professions in the territory where the NPI evaluation study is conducted |
| E2 | Specify the promoter, manager and person responsible for the NPI evaluation study |
| E3 | Declare the competing interests of the NPI evaluation study |
| E4 | Obtain agreement from an ethics committee before conducting the NPI evaluation study |
| E5 | Protect the confidentiality of the data collected on individuals |
| E6 | Use international scientific literature to justify the NPI study |
| E7 | Register as a researcher on the international ORCID registry |
| E8 | Respect international rules of scientific integrity |
| E9 | Systematically publish the results of the NPI evaluation study in a peer-reviewed scientific journal and/or in an open scientific archive |
| E10 | Archive raw data while respecting the confidentiality of personal data |
| E11 | Archive analyzed data and make them accessible for publication while protecting the confidentiality of personal data |
| E12 | Archive the study analysis report |
| E13 | Involve healthcare users concerned by the subject of the study (or their representatives) in the design of the study protocol, the implementation of the study, and the promotion of the results |
| E14 | Present the results to each study participant in an intelligible and systematic manner |
| OP1 | Specify the demographic, medical, and socio-cultural characteristics of the study population |
| OP2 | Identify the relevant experience of traditional or complementary practices in study participants |
| OP3 | Specify the relevant past and current medical treatments that may have significant effects in study participants |
| OI4 | Identify the characteristics of non-pharmacological practices |
| OI4 | Identify the characteristics of non-pharmacological practices |
| OC5 | Use a sufficiently long monitoring time and data collection frequency to assess the effects of the NPI being evaluated on the criteria considered. |
| OO6 | Systematically record health markers (state of health, autonomy, quality of life, survival), and where possible, social, economic and environmental indicators |
| MP1 | Accurately describe the study population and recruitment procedures |
| MP2 | Describe the reasons justifying participant withdrawal from the NPI evaluation study |
| MI3 | Describe the content and context of the hypothetical NPI being evaluated as accurately as possible |
| MI4 | Describe the experience and qualification of the person implementing the hypothetical NPI if necessary |
| MC5 | Describe, as accurately as possible, the experimental condition whose aim is to isolate the mechanism(s) of action studied. |
| MO6 | Analyze the phenomenon observed using scientifically validated tools |
| PP1 | Target a population which may potentially respond to (i.e., be affected by) the NPI prototype |
| PP2 | Justify the number of people needed to answer the research question |
| PP3 | Take into account the past experience of the people participating in the NPI prototype evaluation study |
| PI4 | Describe as accurately as possible the content and context of the NPI prototype |
| PC5 | Define and justify the temporality of the data collected |
| PC6 | Promote the use of a mixed-methods approach |
| PO7 | Collect data on user experience |
| PO8 | Collect data on the experience of the practitioner implementing the NPI prototype |
| PO9 | Define in advance the main health outcome which the NPI prototype is supposed to improve |
| CP1 | Specify the demographic, socioeconomic and cultural characteristics of the population studied |
| CP2 | Specify the medical characteristics of the study participants |
| CP3 | Specify the recruitment strategies used |
| CP4 | Justify the quality of the sampling method |
| CI5 | Name the NPI |
| CI6 | Define the main health objective and the primary outcome |
| CI7 | Describe the content of the NPI |
| CI8 | Describe the psychosocial processes and/or biological mechanisms likely to explain the effect on the main health marker |
| CI9 | Specify the characteristics of the professional(s) implementing the NPI |
| CI10 | Conduct NPI implementation training for all the stakeholders who will implement the NPI during the study |
| CC11 | Conduct a pragmatic controlled intervention study |
| CC12 | Declare the intervention study protocol before its completion on an official platform |
| CC13 | Describe the inclusion and non-inclusion criteria of people participating in the study as well as the exclusion criteria |
| CC14 | Specify secondary objectives |
| CC15 | Justify the choice of the control group |
| CC16 | Guarantee a pragmatic and blind trial |
| CC17 | Always report effectiveness using a statistical test of significance and a confidence interval to report the magnitude of the effect |
| CC18 | Prefer intention-to-treat analyses |
| CC19 | Use resampling techniques as much as possible in statistical evaluation |
| CC20 | When resampling cannot be used, always indicate that the characteristics of the study population align with the assumptions of the parametric model being used |
| CC21 | Check the hypotheses of the *a posteriori* study power calculation, and interpret the significance of the results based on this new calculation |
| CO22 | Choose relevant outcomes measured by validated and sensitive tools |
| CO23 | Specify study withdrawals |
| CO24 | Specify patient compliance to the NPI |
| CO25 | Record concomitant treatments |
| CO26 | Identify adverse events |
| CO27 | Identify unexpected events |
| CO28 | Measure economic indicators as much as possible |
| IP1 | Identify and describe the healthcare service, establishment or territory studied |
| IP2 | Describe the characteristics of study participants |
| II3 | Build on the NPI specifications established during the original intervention study. Detail each NPI used and describe its “invariants” and its “modular components” |
| II4 | Limit the participation of the researchers/evaluators on the study site |
| II5 | Describe the professionals implementing the NPI |
| IC6 | Specify the objectives of the study |
| IC7 | Justify the sample size |
| IC8 | Describe the implementation strategy used |
| IC9 | Describe the data collection process |
| IC10 | Involve operational partners in the field and involve healthcare users |
| IC11 | Describe adaptation approach to the NPI implementation strategy for optimal use in real-world situations |
| IO12 | Describe the measured variables |
| IO13 | Identify the acceptability, fidelity and feasibility of the NPI in different contexts and over time |
| IO14 | Identify the obstacles and drivers to fostering the routine adoption of the NPI |

At the time of writing, 31 scientific societies and three health authorities have provided support letters for the NPIS Model. The complete list of scientific societies and health authorities is available in Annex B, and the support letters are available on the NPIS Model website.^30^

## Discussion

In the following, we discuss several key aspects of this work.

### Definition and Scope of NPIs

A key challenge we addressed in this work was crafting an NPI definition broad enough to meet the needs of all stakeholders, yet specific enough to allow for a rigorous scientific evaluation.^44^ An NPI is an evidence-based, effective, personalized, non-invasive health prevention or care protocol, registered and supervised by a qualified professional. This definition encompasses personalized bodily, psychosocial, and nutritional practices outlined in a clear protocol aimed at preventing, caring for, or curing a health issue recognized in Western medicine. Where appropriate, an NPI serves to complement medications, surgeries, or medical devices. It leverages multiple systemic and dynamic mechanisms that are rationally explainable and present identified risks. An NPI is not a product, a discipline, a diagnostic procedure, a surgery, a public health strategy, an esoteric practice, a way of life, or an organization.

### Double blind RCT vs the NPIS Model

The double-blind randomized control trial (RCT) is widely regarded as the gold standard for drug evaluation. It is designed to evaluate the specific effect of a drug during a clinical trial, while eliminating other effects, including the placebo effect. ^28,42^ However, adapting this approach to NPIs is problematic, as drugs and NPIs face distinct evaluation challenges. For instance, RCTs are not adapted to evaluate NPIs due to the difficulty in creating a control group and implementing blinding. ^43^ Furthermore, the placebo effect is an integral component of NPIs’ effectiveness. ^7^

NPIs also face unique evaluation requirements, such as describing the practical characteristics of an NPI (addressed by prototypical studies) and conditions for successful deployment in a territory including socio-cultural specificities (explored by implementation studies). ^5^

To address these specific needs, we developed the NPIS Model—an evaluation framework encompassing all aspects necessary to assess NPIs comprehensively. Therefore, the NPIS Model goes far beyond a mere modification of the RCT standard by proposing a scientifically sound evaluation framework tailored to the unique nature of NPIs.

### NPIS Model

This work is unique; it gathered more than 1,000 researchers, healthcare practitioners, health operators, and healthcare users to reach a consensus on a rigorous framework—the NPIS Model—to scientifically evaluate NPIs. The outcome is 14 ethical and 63 methodological recommendation items, endorsed by three health authorities and 31 scientific and medical societies. The NPIS Model supports the design, implementation, and dissemination of scientific studies evaluating NPIs, fostering the consolidation of convergent evidence, and promoting best practices in the NPI design and implementation.^5^

The NPIS Model advises that mechanistic studies assess multiple processes simultaneously. It also recommends developing prototypical studies with mixed methods to provide accurate description of intervention protocols,^21,28^ rather than limiting evaluation to overly broad approach (e.g., psychotherapy) or overly narrow components (e.g., a specific manual technique, food ingredient, or equipment). Furthermore, the model recommends pragmatic intervention studies on NPIs,^31^ employing intention-to-treat analyses and examining both biological and self-reported markers of effectiveness, along with adverse effects.^32,33^ It also fosters the use of modern, state of the art statistics methods, such as systematically combining statistical tests and confidence intervals^34,35^ or employing non-parametric resampling techniques, which are well-suited for data collected during intervention studies with complex or unknown distribution.^36,37^ Implementation studies are also encouraged to optimize the real-world dissemination of NPIs, taking into account cultural norms and individual preferences. Finally, the model emphasizes ethical and scientific integrity for both researchers and study promoters.

### Impact of the NPIS Model

Recognition of the NPIS Model by regulatory authorities could foster transparency, methodological rigor, ethical standards, and transferability of NPI research, increasing its value for researchers, healthcare practitioners, healthcare trainers, and study participants. The support from 31 scientific societies and three health authorities underscores the model’s relevance and the need for a structured approach to studying NPIs.

From an economic perspective, recognition of the NPIS Model by relevant authorities could support returns on research investments, create local jobs to manage NPIs, fund effective practices for health insurance and social systems, and reduce avoidable healthcare and hospitalization costs.

From a societal perspective, recognition of the NPIS Model by relevant authorities could enhance information dissemination for the general public, promote sustainable user engagement in safer practices, ensure traceability of NPIs within personalized healthcare pathways, reduce health inequalities, and standardize practices among healthcare practitioners.^13,16,18,32^ Furthermore, accumulating publications of standardized studies would facilitate the identification and comparison of NPIs.^33^ In October 2024, we established a registry of NPIs, accessible to researchers, healthcare practitioners, and the general public.^1^ Each labeled NPI is assigned a unique code to enhance traceability, particularly for large-scale data analyses. Table C1 in Annex C describes the characteristics used to register an NPI.

### How to apply the NPIS Model

A list of 77 items may seem overwhelming at first. However, the NPIS Model should not be viewed as a monolithic framework. Instead, it is structured to include ethical items applicable to all types of studies, along with methodological items tailored to five different types of studies.

Researchers are required to apply all 14 ethical items, in addition to the methodological items relevant to their specific study type. There are 6 methodological items for observational studies, 6 for mechanistic studies, 9 for prototypical studies, 28 for intervention studies, and 14 for implementation studies. For example, a researcher conducting a prototypical study must adhere to a total of 23 items: 14 ethical items and 9 methodological items.

Clinicians can use studies conducted under the NPIS Model to enhance their critical evaluation of NPIs, ensuring they are well-described, explainable, effective, safe, and implementable. Additionally, the NPIS Model helps both researchers and clinicians identify gaps that require further study to safely and efficiently implement an NPI. This process is greatly facilitated by the NPI registry, which centralizes studies following the NPIS Model for labeled NPIs.^1^

### Limitations

One limitation is the absence of a formal vote on the NPI definition. This definition was drafted by 70 contributors in step 1 and reached consensus among 300 contributors in step 2 but was not subject to voting in step 3. Given that no recommendation item validated by consensus in step 2 was rejected by vote in step 3, we believe the consensus on the definition was strong enough to establish the definition’s legitimacy. However, a formal vote would have further reinforced its validity.

Another limitation is that the process was restricted to francophone contributors. We prioritized inclusion across sectors and disciplines over broader international representation, as our existing networks within the francophone community enabled a larger participation of all stakeholders and strong consensus-building. Since our recommendations are grounded in internationally recognized standards and were validated through consensus in step 2 and 3, we do not anticipate significant opposition to the methodological recommendations from the international community. However, some ethical recommendations are based on European standards, and perspectives on regulation and data protection may vary in other regions. Future work should expend this ethical discussion to a broader international audience.

### Next steps

The NPIS Model aims to improve the evaluation of NPIs by providing a comprehensive framework across five study types. To further this objective, we are pursuing three ongoing initiatives. First, we are collaborating with international regulatory bodies to gather feedback and work toward integrating the NPIS Model into regulatory frameworks, similar to the standards governing drug approval. We have already presented the NPIS Model to the European Commission and the WHO, receiving highly positive feedback. Second, we have secured the support of 31 francophone scientific societies and are engaging with their international counterparts to seek additional feedback and endorsement. Lastly, we are advocating for the registration of NPIs evaluated using the NPIS Model in the NPI registry.^1^ This involves presenting the NPIS Model and the registry to health organizations and at NPI-related conferences, with the goal of establishing the NPI registry as the leading internationally recognized registry for NPIs.

## Conclusion

During a two-year consensus effort mobilizing more than 1,000 stakeholders, we developed a standardized framework for evaluating NPIs. Such a framework has many benefits: i) it harmonizes and clarifies epistemological, methodological, and ethical expectations of NPI evaluation studies for researchers; ii) it ensures greater transferability of study results to real-world clinical use of the relevant NPI; iii) it guarantees that programs for professionals in the healthcare, prevention, and social assistance sectors are more operational; iv) it facilitates efficient and safer practices for healthcare users; v) it helps providing decision-makers and regulators with a better understanding of NPIs; vi) it ensures more traceable interventions for health operators; vii) it provides solutions that can be better integrated into the financing strategies of insurance and social solidarity systems. We believe that the NPIS Model can serve as a cornerstone for structuring research on NPIs, fostering dissemination, and ultimately, improving public health policies.

## Data Availability

The anonymized votes, along with the code used to analyze the votes, is publicly available on https://github.com/arnaudlegout/npis_model_stats/

## Contributors (CRediT)

Contributors are given in alphabetical order. Conceptualization: AG, ED, GN

Data curation: AL, GN Formal analysis: AL, PP Funding acquisition: ED, GN

Investigation: all 22 authors

Methodology: AG, AL, BF, ED, FC, FP, GN, NM, PC, PDM, PP

Project administration: AG, ED, GN Resources: AL, ED, GN

Software: AL Supervision: ED, GN

Validation: AL, AG, AMF, ED, GN, PP

Visualization: AL, AG, AMF, ED, GN, PP

Writing – original draft: AL, AG, AMF, ED, GA, GN, PP

Writing – review & editing: AL, AG, AMF, ED, GN, PP

## Data sharing statement

The anonymized votes, along with the code used to analyze the votes and generate Figure 3, is publicly available on https://github.com/arnaudlegout/npis_model_stats/.

## Acknowledgments

Sylvain Agier, Jean-Pierre Aquino, Didier Armaingaud, Patrick Baqué, Yannick Bardie, Caroline Barry, Pierre Louis Bernard, Sylvie Bidon, Dominique Bonneau, Remy Boussageon, Mathis Brier, Philippe Brissaud, Kevin Charras, Antoine Courivaud, Xavier De La Tribonnière, Pascal Demoly, Jacques Desplan, Helene Esperou, Celine Féger, Gianni Franco, David Giovannuzzi, Stephane Guétin, Christian Hervé, Laure Jouatel, Jacques Kopferschmitt, Pierre-Luc L’Hermite, Karen Lambert-Cordillac, Sophie Lantheaume, Emilie Lobertreau, Jean-Bernard Mabire, Herve Maisonneuve, Agnès Mazic de Sonis, Eric Mener, Robert Meslé, Mathilde Minet, Véronique Mondain, Herve Platel, Christian Préfaut, Henri Pujol, Stephanie Ranque-Garnier, Sylvie Rapior, Carole Robert, Thierry Schaeverbeke, Alain Segu, Laurent Stubbe, Christine Tabuenca, Henri Truong Tan Trung, Marion Trousselard, Alain Warnery, Aline Weber

## Supplemental material

Supplementary material can be found in Appendix A to F.

## Declaration of generative AI and AI-assisted technologies in the writing process

During the preparation of this work the authors used ChatGPT to improve language. The use was limited to sentence fragments. All propositions were carefully reviewed, modified, and integrated by the authors. We never used a generative AI to analyze results and write full sentences or paragraphs. The authors take full responsibility for the content of the publication.

