## Supplementary material for "The NPIS Model: A Standardized, Consensus-Based Framework for Evaluating Non-Pharmacological Interventions"

### Annex A

**NPIS Model, a Standardized Scientific and Ethical Evaluation Framework of Non-Pharmacological Interventions (NPIs) in the Domain of Health**

*Ethical recommendations*

| Code | Ethical Items | Explanation |
|---|---|---|
| E1 | **Respect the laws, regulations and ethics charters of the research professions in the territory where the NPI evaluation study is conducted** | All NPI evaluation studies must comply with the law on research involving humans in their country. This legal framework applies to principal investigators, persons associated with the study, persons participating in the study, the study sponsor, and the investigative center. As an example, for studies performed in Europe, an NPI evaluation study must not fall under European Regulation 536/2014 relating to clinical trials of medicinal products for human use, European Regulation 2017/745 relating to medical devices, or European Regulation 2283/2015 relating to food supplements. |
| E2 | **Specify the promoter, manager and person responsible for the NPI evaluation study** | Specify the organization and person responsible for the study, particularly for insurance and legal issues. |
| E3 | **Declare the competing interests of the NPI evaluation study** | Indicate the competing interests of the study for all oral or written communication for a period of 5 years. Furthermore, specify all the kinds of support received |
| E4 | **Obtain agreement from an ethics committee before conducting the NPI evaluation study** | Submit the study protocol to a research ethics committee. Agreement from an ethics committee is required both to commence the study and for all its stages until its publication. The protocol can be subject to *a posteriori* control. |
| E5 | **Protect the confidentiality of the data collected on individuals** | Comply with the data protection principles of your country. For instance, for European studies, or studies involving European participants, the study must comply to the European Union's General Data Protection Regulation. |
| E6 | **Use international scientific literature to justify the NPI study** | Consult general health databases (e.g., Pubmed, Cochrane, Science Direct, Google Scholar, HAL, CORE), and databases specializing in NPIs (e.g., PEDro, APA PsycInfo). |
| E7 | **Register as a researcher on the international ORCID registry** | Register on the Open Researcher and Contributor ID (ORCID) registry. Scientific journals require this individual code to publish a study and facilitate traceability of the researcher. |
| E8 | **Respect international rules of scientific integrity** | Irrespective of the protocol for the NPI evaluation study, follow the principles and obligations of the Singapore Declaration on Research Integrity.[1] |
| E9 | **Systematically publish the results of the NPI evaluation study in a peer- reviewed scientific journal and/or in an open scientific archive** | Publish the results of the study, whether positive or negative. Consult the list of peer-reviewed scientific health journals in SCImago. |
| E10 | **Archive raw data while respecting the confidentiality of personal data** | Making raw data accessible enables their reuse for new analyses, ancillary studies and meta-analyses. Guarantee the sustainability of these data. |
| E11 | **Archive analyzed data and make them accessible for publication** | Ensuring the accessibility of analyzed data enables their reuse for new analyses, ancillary studies, and meta-analyses. |

| | | |
|---|---|---|
| | **while protecting the confidentiality of personal data** | Guarantee the sustainability of these data. Specify if, where, and how the data are accessible. |
| E12 | **Archive the study analysis report** | Ensuring access to the complete data analysis report encourages interdisciplinary views, which are particularly relevant in the study of NPIs. |
| E13 | **Involve healthcare users concerned by the subject of the study (or their representatives) in the design of the study protocol, the implementation of the study, and the promotion of the results** | In all stages of the study, involve participants who directly benefit from it (e.g., patients, associations) in its design and implementation. |
| E14 | **Present the results to each study participant in an intelligible and systematic manner** | Adapt the format of the presentation of the results according to the levels of education, culture and knowledge of the study participants. |

***Methodological Recommendations***

<u>Observational study</u>

In an observational study on humans, researchers do not intervene during events and only observe a non-pharmacological practice. It could be an approach, a method, a technique, or an ingredient. This is done either prospectively (e.g., cohort) or retrospectively (e.g., datamining, big data analysis). In 2007, the *Enhancing the QUAlity and Transparency Of health Research* network established an international recommendation for reporting observational studies in epidemiology, named STROBE.[2] STROBE details how the results of a study should be presented in a scientific article (title, abstract, introduction, method, results, discussion, and other necessary information).

Population

| Code | Methodological Items | Explanation |
|---|---|---|
| OP1 | **Specify the demographic, medical, and socio-cultural characteristics of the study population** | Collecting data (at the very least) on study participants' age, gender, profession, and place of residence helps researchers to identify NPI responders and limit population biases. |
| OP2 | **Identify the relevant experience of traditional or complementary practices in study participants** | Data collection on traditional or complementary practices habits provides relevant information on patients' expectations about the possible effects of the NPI. |
| OP3 | **Specify the relevant past and current medical treatments that may have significant effects in study participants** | Data collection on biomedical treatments is necessary to consider the influence of these treatments on the effects observed. |

Intervention

| Code | Methodological Items | Explanation |
|---|---|---|
| OI4 | **Identify the characteristics of non-pharmacological practices** | The characterization of a hypothetical NPI requires the description of its content (e.g., number, duration and frequency of sessions, mode of use of the equipment, place of practice, practitioner), the NPI access conditions (i.e., face-to-face or telemedicine), and the description of its components (e.g., equipment, technique, skill, ingredient). Two or more NPIs may be combined. |

Comparison

| Code | Methodological Items | Explanation |
|---|---|---|
| OC5 | **Use a sufficiently long monitoring time and data collection frequency to assess the effects of the NPI being evaluated on the criteria considered.** | NPIs rarely have immediate effects on health. A sufficiently long monitoring time with sufficient data collection frequency is required to observe the kinetics of the different markers evaluated. |

Outcome

| Code | Methodological Items | Explanation |
|---|---|---|
| OO6 | **Systematically record health markers (state of health, autonomy, quality of life, survival), and where** | An analysis of health data (e.g., benefits, adverse effects), autonomy (e.g., behaviors), quality of life (e.g., patient-reported outcomes) and life expectancy (e.g., life expectancy without loss of quality of life), as well as social (e.g., social participation), medico-economic (e.g., hospitalization, work |

| | | |
|---|---|---|
| | **possible, social, economic and environmental indicators** | stoppage) and environmental (e.g., energy expenditure) analyses, enable the identification of possible systemic effects of an NPI on a cohort. |

Mechanistic study

In a mechanistic study, researchers highlight the biological mechanisms and active psychosocial processes and interactions with the environment (e.g., exposome), which explain the benefits of the NPI for health, autonomy, quality of life, or survival.

Population

| Code | Methodological Items | Explanation |
|---|---|---|
| MP1 | **Accurately describe the study population and recruitment procedures** | This type of study makes it possible to isolate the mechanisms at play (e.g., active principle in biology, processes in human science), which explain the effect of an NPI on health. Furthermore, the study population must be described accurately. Depending on the question asked, the data obtained can be compared to control situations. |
| MP2 | **Describe the reasons justifying participant withdrawal from the NPI evaluation study** | Study participants may withdraw their consent, be excluded because of protocol violation, be lost to follow-up, experience a side effect of the NPI, or declare a contraindication. |

Intervention

| Code | Methodological Items | Explanation |
|---|---|---|
| MI3 | **Describe the content and context of the hypothetical NPI being evaluated as accurately as possible** | This description makes it possible to consider the effect of the context on the mechanism(s) studied. |
| MI4 | **Describe the experience and qualification of the person implementing the hypothetical NPI if necessary** | This description makes it possible to consider the effect of the practitioner's experience on the mechanism(s) studied. |

Comparison

| Code | Methodological Items | Explanation |
|---|---|---|
| MC5 | **Describe, as accurately as possible, the experimental condition whose aim is to isolate the mechanism(s) of action studied.** | The study design highlights the mechanism(s) of action and the process(es). A mechanism can impact several markers. Whether a study targets the microscopic or macroscopic level, the researcher must be aware that an NPI mobilizes several mechanisms simultaneously. The method of measuring the observed phenomenon must be reproducible. |

Outcome

| Code | Methodological Items | Explanation |
|---|---|---|
| MO6 | **Analyze the phenomenon observed using scientifically validated tools** | An NPI mobilizes mechanisms and processes that can be observed on biological, physiological, behavioral, psychological, and social markers. |

Prototypical Study

In a prototypical study, researchers identify all the practical characteristics of an NPI by using methods for collecting information on practitioners and on user experience. The empirical study details the NPI protocol through feedback from practitioners and target users. The NPI prototype is then described along the grounds of the NPIS Model (designation, main health benefit, secondary benefits, risks, mechanisms, target population, protocol, professional, context of use) and is recorded in a sort of user manual intended for professionals in the health field. It details the contents of the NPI, the target population, the professional prerequisites to implement it, and the different contexts where the NPI can be used, to guarantee the reproducibility of its effects on health markers.

Population

| Code | Methodological Items | Explanation |
|---|---|---|
| PP1 | **Target a population which may potentially respond to (i.e., be affected by) the NPI prototype** | An NPI cannot benefit everyone in the same way. The NPI evaluation study must target a homogeneous population with the objective of improving this population's state of health. |
| PP2 | **Justify the number of people needed to answer the research question** | Having a minimum number of people participating in the study makes it possible to consolidate the reproducibility of the NPI. |
| PP3 | **Take into account the past experience of the people participating in the NPI prototype evaluation study** | The effect of an NPI may differ depending on a person's past experiences. |

Intervention

| Code | Methodological Items | Explanation |
|---|---|---|
| PI4 | **Describe as accurately as possible the content and context of the NPI prototype** | The NPI evaluation study makes it possible to design the NPI prototype with an original name that describes its content and its implementation conditions. Doing this differentiates the NPI from an approach or a component. The NPI is therefore characterized, described, and deployed to become reproducible in a similar context. |

Comparison

| Code | Methodological Items | Explanation |
|---|---|---|
| PC5 | **Define and justify the temporality of the data collected** | The evaluation of the NPI prototype can take place at any of the following stages: before, during, or after its implementation. Furthermore, the evaluation can be repeated. |
| PC6 | **Promote the use of a mixed-methods approach** | A methodology which collects qualitative and quantitative data is advantageous to collect the multiple impacts of an NPI. |

Outcome

| Code | Methodological Items | Explanation |
|---|---|---|
| PO7 | **Collect data on user experience** | The study should make it possible to clarify the satisfaction, acceptability, and level of support for the NPI (disincentives and motivations). |
| PO8 | **Collect data on the experience of the practitioner implementing the NPI prototype** | The study should make it possible to specify the conditions for the routine implementation of the NPI and the resources required. |
| PO9 | **Define in advance the main health outcome which the NPI** | The study must specify the main health criterion targeted by the NPI and, if possible, its secondary criteria. These criteria may be unique or composite. |

| | |
|---|---|
| | **prototype is supposed to improve** |

Intervention study

In a clinical trial with patients or an intervention study with people without a declared disease, researchers highlight the level of effectiveness of an NPI on a target population, that is the benefits and risks on this population's health. The study focuses on establishing whether there is a direct causal relationship between the NPI and its health effects. This method provides the best evidence that under similar conditions, the NPI will provide the same health benefits and cause the same side effects and health risks. Researchers must use the SPIRIT guide to communicate the results of a clinical trial.[3,4] Furthermore, researchers must use the TIDieR guide[5] to describe the intervention, so that it can be better replicated in health practice or research.[6] Moreover, researchers must use the CONSORT Nonpharmacologic Treatments guide for randomized trials.[7]

Population

| Code | Methodological Items | Explanation |
|---|---|---|
| CP1 | **Specify the demographic, socioeconomic and cultural characteristics of the population studied** | Providing at least the following population characteristics - age, gender, and at least one socioeconomic indicator - makes it possible to specify which populations may potentially respond to (i.e., be affected by) the NPI being evaluated, and to promote the comparability and reproducibility of the study. The characteristics of people not included in the study should also be specified. |
| CP2 | **Specify the medical characteristics of the study participants** | The nature and severity of participants' pathologies, risk factors, and medical history may modify the observed effects of the NPI. Collecting information on biomedical treatments is necessary to consider their influence on the observed effects. |
| CP3 | **Specify the recruitment strategies used** | The recruitment context influences the observed effects. It is important to specify whether participants in the study received financial compensation. |
| CP4 | **Justify the quality of the sampling method** | Describe how the sampling method used is representative of the target population, outline the process of how sampling was conducted, and identify any potential biases. |

Intervention

| Code | Methodological Items | Explanation |
|---|---|---|
| CI5 | **Name the NPI** | The study must explicitly cite the name of the NPI, and where applicable, its acronym and the persons who designed it. |
| CI6 | **Define the main health objective and the primary outcome** | The study must confirm an hypothesis for the effect of the NPI on a main health marker (e.g., risk behavior, symptom, sequelae, disease, functional capacity, survival, quality of life) – also called the primary outcome – with a defined action (prevent, care, or cure). The study must determine the specific effect, the overall effect, and the contextual effect of the NPI evaluated. |
| CI7 | **Describe the content of the NPI** | The study must describe the NPI, its components (e.g., ingredients, techniques, skills), its procedure (e.g., sessions, dose/intensity, duration, frequency), and the equipment required to make it reproducible. The conditions of access to the intervention and possible interactions with biomedical treatments must also be specified (e.g., medical prescription). |
| CI8 | **Describe the psychosocial processes and/or biological** | Develop a rationale describing the principles of actions that may explain the expected benefits of the NPI. |

| | | |
|---|---|---|
| | **mechanisms likely to explain the effect on the main health marker** | |
| CI9 | **Specify the characteristics of the professional(s) implementing the NPI** | Name the job of the professional implementing the NPI and describe their skills and qualifications. |
| CI10 | **Conduct NPI implementation training for all the stakeholders who will implement the NPI during the study** | This involves guaranteeing homogeneity and ensuring the standardization of practice between groups, or between establishments collaborating in the study. |

Comparison

| Code | Methodological Items | Explanation |
|---|---|---|
| CC11 | **Conduct a pragmatic controlled intervention study** | The study evaluates the real-world effectiveness of the NPI. The study is intended to isolate the specific effect of the NPI on the main health outcome. The choice of comparison groups and the method of assigning people to groups must be justified. |
| CC12 | **Declare the intervention study protocol before its completion on an official platform** | Several reporting platforms exist upstream of the intervention study protocol. The most used general platform is Clinical Trials. An example of a platform specialized in physiotherapy is PEDro. |
| CC13 | **Describe the inclusion and non-inclusion criteria of people participating in the study as well as the exclusion criteria** | Justify the criteria and the number of persons needed to treat. |
| CC14 | **Specify secondary objectives** | Detail all the health criteria likely to be modified by the NPI being evaluated. |
| CC15 | **Justify the choice of the control group** | The control group must make it possible to evaluate the specific effect of the NPI being tested. |
| CC16 | **Guarantee a pragmatic and blind trial** | The possibility of blinding must take precedence over the difficulty in implementing the NPI. The hypothesis to which each group is blinded, including the evaluator, must be defined. The professional who implements the NPI cannot always be blinded. The people participating in the trial should be blinded as much as possible. Evaluators should be blinded as much as possible. In all cases, specify the measures taken to ensure blinding. |
| CC17 | **Always report effectiveness using a statistical test of significance and a confidence interval to report the magnitude of the effect** | Always combine the confidence interval, p-value, and effect size of all the outcomes assessed. |
| CC18 | **Prefer intention-to-treat analyses** | Intention-to-treat analyses are closer to real life and are applied in the field of health. Include an analysis with imputation of missing data either in the main analysis or in a sensitivity analysis. |
| CC19 | **Use resampling techniques as much as possible in statistical evaluation** | Resampling techniques (permutation test, bootstrap) are more robust than parametric statistical tests in most cases. As they are also simpler to implement and easier to interpret, they should always be preferred. |
| CC20 | **When resampling cannot be used, always indicate that the characteristics of the study population align with the** | Resampling is not suitable for small samples or samples not randomly chosen from the target population. In this case, a parametric model can give valuable results if - and only if - the characteristics of the study population align with the model |

| | | |
|---|---|---|
| | **assumptions of the parametric model being used** | assumptions. One must always check for this and report that it is indeed the case. |
| CC21 | **Check the hypotheses of the *a posteriori* study power calculation, and interpret the significance of the results based on this new calculation** | The calculation of the study power is useful to provide information on the reason for the non-significance of a result (e.g., number of people participating in the study is too low *a posteriori*). It can help refine hypotheses for calculating the study power and the minimum number of people needed to participate in a future study. |

Outcome

| Code | Methodological Items | Explanation |
|---|---|---|
| CO22 | **Choose relevant outcomes measured by validated and sensitive tools** | Use objective and subjective criteria (e.g., patient-reported outcomes) employing the SMART approach (Specific, Measurable, Achievable, Realistic, and Timely), measured with validated instruments in the local language and, if possible, with a minimal clinically important difference (MCID). |
| CO23 | **Specify study withdrawals** | Indicate the withdrawal rates and reasons, as well as the rates of loss to follow-up. Limit the exit of people participating in the study irrespective of the group (i.e., intervention group, control group), even in the event of withdrawal. |
| CO24 | **Specify patient compliance to the NPI** | Measure the patient compliance rate (percentage of completion of scheduled sessions). |
| CO25 | **Record concomitant treatments** | Other NPIs, medicine, surgery, medical devices, hospital admission, etc. |
| CO26 | **Identify adverse events** | Healthcare practices involve risks. Ensure the research team has the means to search for adverse events as part of a vigilance system and report them in the presentation of results. |
| CO27 | **Identify unexpected events** | An intervention study/clinical trial may reveal unexpected health benefits. Record observations of the professionals implementing the NPI and of participants (or their care givers). |
| CO28 | **Measure economic indicators as much as possible** | NPIs can impact direct expenses (e.g., the NPI itself, biomedical treatment, care, hospitalization) and indirect (e.g., sick leave, caregiver contributions) expenses. |

Implementation study

In implementation studies, researchers determine the conditions for successful deployment of an NPI within a specific territory and identify modalities for adjusting it to various context (e.g., territorial, social, cultural, economic). Such studies provide specifications for transferability and usage precautions, allowing field-based teams to make necessary adjustment without compromising the NPI's effectiveness on health markers demonstrated in previous intervention studies. Implementation studies also formalize the traceability procedures or the elements of quality improvement. An international recommendation for reporting implementation studies, named STaRI, was established in 2017.[8] Depending on existing knowledge about the implementation context and potential deployment strategies, implementation studies may focus on identifying barriers and facilitators, developing or selecting implementation strategies, and even comparing the value of different implementation strategies. This approach is particularly useful for understanding the adoption, effective implementation, and sustainability of NPIs within their intended context.

Population

| Code | *Methodological Items* | *Explanation* |
| --- | --- | --- |
| IP1 | **Identify and describe the healthcare service, establishment or territory studied** | Describe the meso- and macro-environmental characteristics of the healthcare service, establishment or territory targeted for the implementation of the NPI (social, economic, political, organizational, cultural and structural specificities). This makes it possible to estimate the external validity of the study. In addition, the modification of these characteristics can influence the implementation of the NPI over time and produce unpredictable effects which will require adaptation. |
| IP2 | **Describe the characteristics of study participants** | Describe the eligibility criteria for study participants. The description provides information on the possibility of implementing the NPI in similar populations. |

Intervention

| Code | Methodological Items | Explanation |
| --- | --- | --- |
| II3 | **Build on the NPI specifications established during the original intervention study. Detail each NPI used and describe its "invariants" and its "modular components"** | The "invariants" are the essential and indispensable elements of the NPI. In contrast, "modular components" are elements, structures and systems that can be adapted depending on the location of the study and the users, without compromising the integrity of the NPI. Insufficient adherence to the invariants can dilute the effect of the NPI, whereas insufficient adaptation of the "modular components" can inhibit its effect. |
| II4 | **Limit the participation of the researchers/evaluators on the study site** | This provision consolidates the validity of the study. The researchers must limit personal involvement, from data collection to the training of the professionals who will implement the NPI. If the researchers cannot limit their involvement, justification is required. |
| II5 | **Describe the professionals implementing the NPI** | Describe the qualifications, roles, and training of the professionals implementing each NPI and the number of professionals implementing it. |

Comparison

| Code | Methodological Items | Explanation |
| --- | --- | --- |
| IC6 | **Specify the objectives of the study** | Describe the objectives of the implementation of the NPI (e.g., acceptability, adoption, commitment, safety, scope, sustainability, transferability, integration into the care/health pathway, cost). |
| IC7 | **Justify the sample size** | Justify the sample size according to the constraints of the study (budgetary, practical, data analysis). Depending on the design and objectives of the study, a sample size calculation is possible. |
| IC8 | **Describe the implementation strategy used** | Describe how the NPI is implemented to enable its adoption, transferability, and sustainability. |
| IC9 | **Describe the data collection process** | The data collection process concerns the extraction of routine clinical data and risk assessment data (side effects, interactions). It is recommended to create a standardized recording procedure to avoid inconsistencies in entries (e.g., missing data, under- or over-estimation). |
| IC10 | **Involve operational partners in the field and involve healthcare users** | Involve operational partners in the field and users of the NPI from the conception of the protocol all the way to the analysis of results. Develop a formal implementation strategy together |

| | | |
|---|---|---|
| | | that overcomes obstacles and empowers facilitators to increase adoption of the intervention. |
| IC11 | **Describe adaptation approach to the NPI implementation strategy for optimal use in real-world situations** | The adaptation of the NPI implementation strategy must be described. The complexity of the implementation context — inherent to the heterogeneity and the needs of the study population — will necessarily require the implementation strategy to be adapted (e.g., refresher training courses for persons implementing the NPI to maintain their commitment to it). Social aid strategies to compensate for social inequalities must be clarified (e.g., compensation for travel costs for health consultations). |

Outcome

| Code | Methodological Items | Explanation |
|---|---|---|
| IO12 | **Describe the measured variables** | Describe the health and social contexts, and, if possible, the political context in which data collection will occur. |
| IO13 | **Identify the acceptability, fidelity and feasibility of the NPI in different contexts and over time** | Fidelity is the most important element for the successful implementation of an NPI. Evaluate acceptability, fidelity, and feasibility iteratively to increase the chances of transferability and sustainability of the NPI in a real-world context (through adaptations), and to evaluate the impact of the implementation. It is preferable to consider these "implementability" factors when developing the study. |
| IO14 | **Identify the obstacles and drivers to fostering the routine adoption of the NPI** | This evaluation must be conducted with all the stakeholders involved (e.g., people participating in the study, establishment, organization, promoter, decision-makers). |

#### Annex B

**Scientific societies supporting the NPIS Model (31)**

Association des Chercheurs en Activités Physiques et Sportives (ACAPS)
Association Française d'Urologie (AFU)
Association Française de Psychiatrie Biologique et Neuropsychopharmacologie (AFPBN)
Association Francophone des Soins Oncologiques de Support (AFSOS)
Collège de la Médecine Générale (CMG)
Collège National des Généralistes Enseignants (CNGE)
Collège National des Sages-femmes de France (CNSF)
Société d'Éducation Thérapeutique Européenne (SETE)
Société de Pneumologie de Langue Française (SPLF)
Société Française d'Accompagnement et de Soins Palliatifs (SFAP)
Société Française d'Alcoologie (SFA)
Société Française d'Allergologie (SFA)
Société Française d'Anesthésie Réanimation (SFAR)
Société Française d'Endocrinologie (SFE)
Société Française d'Étude et Traitement de la Douleur (SFETD)
Société Française de Cardiologie (SFC)
Société Française de Neurologie (SFN)
Société Française de Nutrition (SFN)
Société Française de Pédiatrie (SFP)
Société Française de Physiothérapie (SFP)
Société Française de Psychiatrie de l'Enfant et de l'Adolescent et Disciplines Associées (SFPEADA)
Société Française de Psychologie (SFP)

Société Française de Rhumatologie (SFR)
Société Française de Santé Publique (SFSP)
Société Française de Tabacologie (SFT)
Société Française et Francophone d'Éthique Médicale (SFFEM)
Société Francophone d'Étude et de Recherche en Orthoptie (SFERO)
Société Francophone de Néphrologie, Dialyse et Transplantation (SFNDT)
Société Francophone de Santé et Environnement (SFSE)
Société Francophone Nutrition Clinique et Métabolisme (SFNCM)
Société Nationale Française de Gastro-Entérologie (SNFGE)

**Authorities supporting the NPIS Model (3)**

Centre National des Soins Palliatifs et de la Fin de Vie
Institut National du Cancer (INCA)
National Clinical Research Infrastructure F-CRIN

#### Annex C

*Table C1: Descriptive characteristics of an NPI. The column "Study type" represents the required type of studies published in a peer-reviewed scientific journal to validate the corresponding characteristic of the NPI. In this column: O represents an observational study, M a mechanistic study, P a prototypical study, C a clinical study, and I an implementation study.*

| Characteristic | Description | Study type |
|---|---|---|
| Designation | Name (abbreviation if applicable). | P, C |
| Main health benefit | Health problem prevented, cared, or cured. | C |
| Secondary benefits | Benefits for other health markers (biological and/or psychosocial). | C, I |
| Risks | Side effect(s), at-risk interaction(s). | O, M, C, I |
| Mechanisms | Biological mechanism(s) of action, and/or active psychosocial process(es) explaining the benefits for the health markers of interest. | M |
| Target population | Public responder, contraindication(s). | O, P, C, I |
| Protocol | Components (e.g., ingredients, techniques, gestures), procedure (e.g., duration, number and frequency of sessions, dose), equipment (e.g., physical, digital) required to guarantee the reproducibility of the effects on health. | P, C |
| Professional | Required qualifications. | P, C, I |
| Context of use | Places of practice, condition of use (e.g., medical prescription), good implementation practices, precautionary measures, maintenance strategy, regulatory characteristics, initiators. | P, C, I |

#### Annex D

**List of the 22 experts**
Gregory Ninot,[a,b]
Emeline Descamps,[c]
Ghislaine Achalid,[d]
Sébastien Abad,[a,e]
Fabrice Berna,[f,g]

Christine Belhomme,[h]
Pierre Louis Bernard,[i]
François Carbonnel,[a]
Patrizia Carrieri,[j]
Patricia Dargent-Molina,[k]
Frederic Fiteni,[a,l]
Aude-Marie Foucaut,[m]
Alice Guyon,[n]
Arnaud Legout,[o]
Beatrice Lognos,[a]
Nicolas Molinari,[a,p]
Julien Nizard,[q,r]
Michel Nogues,[d]
François Paille,[s,t]
Pierrick Poisbeau,[u]
Lise Rochaix,[v]
Bruno Falissard[w]

One expert is not coauthor of this paper: Pierre Louis Bernard

**Affiliation of the experts**
[a] Desbrest Institute of Epidemiology and Public Health, University of Montpellier, INSERM, Montpellier, France
[b] Montpellier Cancer Institute, Montpellier, France
[c] Inserm Unité ToNIC, UMR 1214, CHU Purpan, Toulouse, France Centre National de la Recherche Scientifique, Toulouse, France
[d] Non-Pharmacological Intervention Society (NPIS), Paris, France
[e] CHU Rouen, Rouen, France
[f] University of Strasbourg, France
[g] Strasbourg University Hospital, Strasbourg, France
[h] Allie Sante, Amboise, France
[i] University of Montpellier, Montpellier, France
[j] INSERM, U912 SESSTIM, University Aix Marseille, IRD, UMR-S912, Marseille, France
[k] Universite Paris Cite et Universite Sorbonne Paris Nord, INSERM, INRAE, Center for Research in Epidemiology and StatisticS (CRESS), 75004 Paris, France
[l] CHU Nimes, Nimes, France
[m] Health Educations and Promotion Laboratory, UR 3412, University Sorbonne Paris North, Bobigny, France
[n] Aix Marseille University, Centre National de la Recherche Scientifique (CNRS), UMR 7770 CRPN (Center of research in Psychology and Neurosciences), Marseille, France
[o] Centre INRIA from University Cote d'Azur, Sophia Antipolis, France
[p] CHU Montpellier, Montpellier, France
[q] EA4391 Nervous and Therapeutics Excitability, Nantes, France
[r] Pain, Palliative and Supportive care, Ethics Department, UIC22, University Hospital, Nantes, France
[s] University of Nancy, Nancy, France
[t] CHU Nancy, France
[u] University of Strasbourg, Centre National de la Recherche Scientifique, Laboratoire des Neurosciences Cognitives et Adaptatives, Strasbourg, France
[v] Paris School of Economics et University of Paris 1, Paris, France Hospinnomics, Hôtel-Dieu, HPHP, Paris, France
[w] Centre for Epidemiology and Population Health (U1018 INSERM), Villejuif, France University Paris Saclay, Le Kremlin-Bicetre, France

**Domain of competencies of the experts**
Biostatistics, biology, economics, ethics, health-applied mathematics and computer science, neuroscience, public health, behavioral sciences, humanities, addictology, geriatrics, pain medicine, general medicine, nutrition, oncology, pediatrics, prevention, psychiatry, rehabilitation, palliative care.

### Annex E

Profession as stated by the 503 participants in the online voting platform.

Healthcare user: 76 voters
Reflexologist: 67 voters
Shiatsu Specialist: 65 voters
Researcher: 58 voters
Doctor: 56 voters
Healthcare Operator: 41 voters
Osteopath: 35 voters
Masseur: 15 voters
Psychologist: 11 voters
Nurse: 11 voters
Sophrologist: 11 voters
Art Therapist: 9 voters
Pharmacist: 8 voters
APA Teacher (Adapted Physical Activity Teacher): 7 voters
Naturopath: 6 voters
Physiotherapist: 4 voters
Healthcare Manager: 3 voters
Fasciatherapist: 3 voters
Hypnotherapist: 3 voters
Music Therapist: 2 voters
Chinese Medicine Practitioner: 2 voters
Psychomotor Therapist: 2 voters
Nursing Assistant: 1 voter
Chiropractor: 1 voter
Life Coach: 1 voter
Dentist: 1 voter
Dietitian: 1 voter
Speech Therapist: 1 voter
Professional: 1 voter
Midwife: 1 voter

### Annex F

We present in this section the two guideline recommendation instruments we followed to build the NPIS Model: AGREE II[10] and the EQUATOR recommendations.[9] We explain shortly for each item when relevant who was in charge of the item and how it has been addressed.

*Table E1: The AGREE II score sheet. Instead of providing a self-assessed score, we explain where we addressed the item.*

| Domain | Item | Where we addressed the item |
|---|---|---|
| | 1. The overall objective(s) of the guideline is (are) specifically described. | Introduction |

| | | |
|---|---|---|
| Scope and purpose | 2. The health question(s) covered by the guideline is (are) specifically described. | Introduction |
| | 3. The population (patients, public, etc.) to whom the guideline is meant to apply is specifically described. | Introduction, NPIS Model in Annex A |
| Stakeholder involvement | 4. The guideline development group includes individuals from all the relevant professional groups. | Step 1, 2, and 3 |
| | 5. The views and preferences of the target population (patients, public, etc.) have been sought. | Step 1, 2, and 3 |
| | 6. The target users of the guideline are clearly defined. | NPIS Model in Annex A |
| Rigor of development | 7. Systematic methods were used to search for evidence. | Research in context, step 1 |
| | 8. The criteria for selecting the evidence are clearly described. | Research in context, step 1 |
| | 9. The strengths and limitations of the body of evidence are clearly described. | Discussion |
| | 10. The methods for formulating the recommendations are clearly described. | Step 1 and 2 |
| | 11. The health benefits, side effects and risks have been considered in formulating the recommendations. | Step 1 and 2 |
| | 12. There is an explicit link between the recommendations and the supporting evidence. | Step 1 and 2 |
| | 13. The guideline has been externally reviewed by experts prior to its publication. | Step 4 |
| | 14. A procedure for updating the guideline is provided. | Future work section |
| Clarity of presentation | 15. The recommendations are specific and unambiguous. | Step 2 |
| | 16. The different options for management of the condition or health issue are clearly presented. | Step 2 |
| | 17. Key recommendations are easily identifiable. | NPIS Model in Annex A |
| Applicability | 18. The guideline describes facilitators and barriers to its application. | NPIS Model in Annex A (Implementation study) |
| | 19. The guideline provides advice and/or tools on how the recommendations can be put into practice. | NPIS Guidelines published in NPIS Web site<br><br>https://npisociety.org/npismodel/ |
| | 20. The potential resource implications of applying the recommendations have been considered. | NPIS Model in Annex A |
| | 21. The guideline presents monitoring and/ or auditing criteria. | NPIS Model in Annex A |
| Editorial independence | 22. The views of the funding body have not influenced the content of the guideline. | Yes |
| | 23. Competing interests of guideline development group members have been recorded and addressed. | Yes |

*Table E2: The EQUATOR recommendation*

| Step | Item number | Identify the need for a guideline | Performed action |
|---|---|---|---|
| Initial steps | 1 | Identify the need for a guideline | Done in Carbonnel et al[11] |
| | 1.1 | Develop new guidance | Done in Carbonnel et al[11] + steering committee |
| | 1.2 | Extend existing guidance | Done in Carbonnel et al[11] + steering committee |
| | 1.3 | Implement existing guidance | Done in Carbonnel et al[11] + steering committee |
| | 2 | Review the literature | Done in Carbonnel et al[11] + step 1 |
| | 2.1 | Identify previous relevant guidance | Done in Carbonnel et al[11] + step 1 |
| | 2.2 | Seek relevant evidence on the quality of reporting in published research articles | Done in Carbonnel et al[11] |
| | 2.3 | Identify key information related to the potential sources of bias in such studies | Done in Carbonnel et al[11] |
| | 3 | Obtain funding for the guideline initiative | INSERM, France |
| Pre-meeting activities | 4 | Identify participants | Steps 1 to 3 |
| | 5 | Conduct a Delphi exercise | Although we did not conduct a formal Delphi exercise, our approach in steps 1, 2, and 3 closely resembled one. We prioritized group discussions to reach a consensus rather than relying on questionnaires. |
| | 6 | Generate a list of items for consideration at the face-to-face meeting | Based on EQUATOR recommendations, |
| | | Prepare for the face-to-face meeting | Steering committee |
| | 7.1 | Decide size and duration of the face-to-face meeting | Steering committee |
| | 7.2 | Develop meeting logistics | NPI Society |
| | 7.3 | Develop meeting agenda | NPI Society |
| | 7.3.1 | Consider presentations on relevant background topics, including summary of evidence | Steering committee |
| | 7.3.2 | Plan to share results of Delphi exercise, if done | We shared with all participants the working and final document. With this article, we intend to share the final results of step 3 to the journal audience. |
| | 7.3.3 | Invite session chairs | Steering committee |
| | 7.4 | Prepare materials to be sent to participants prior to meeting | Steering committee |
| | 7.5 | Arrange to record the meeting | Steering committee |

| | | | |
|---|---|---|---|
| The face-to-face consensus meeting itself | 8 | Present and discuss results of pre-meeting activities and relevant evidence | Steering committee |
| | 8.1 | Discuss the rationale for including items in the checklist | Online meetings step 1 and 2 |
| | 8.2 | Discuss the development of a flow diagram | Not applicable |
| | 8.3 | Discuss strategy for producing documents; identify who will be involved in which activities; discuss authorship | Steering committee |
| | 8.4 | Discuss knowledge translation strategy | Steering committee |
| Post-meeting activities | 9. | Develop the guidance statement | Steering committee |
| | 9.1 | Pilot test the checklist | Expert committee |
| | 10 | Develop an explanatory document (E&E) | NPIS Guidelines published in NPIS Web site<br>https://npisociety.org/npismodel/ |
| | 11 | Develop a publication strategy | Steering and expert committees |
| | 11.1 | Consider multiple and simultaneous publications | Steering committee<br>NPIS Guidelines<br>https://npisociety.org/npismodel/<br>NPIS Model published on the open archive HAL<br>https://hal.science/hal-04360550 |
| Post-publication activities | 12 | Seek and deal with feedback and criticism | Meetings in step 1 and 2<br>Votes in step 3<br>Support from 31 scientific society and 3 health authorities in step 4<br>The NPIS Model has been presented to all French health authorities (the Senate, the Ministry of Health, the French Authority for Health, the French Public Health Agency, the French Academy of Medicine, the General Inspectorate for Social Affairs, the Health Innovation Agency, the French Health Insurance System, the French National Solidarity Fund for Independent Living, and the French Pension Fund). |
| | 13 | Encourage guideline endorsement | NPIS Summit<br>https://npisummit.org/<br>NPI Forum<br>https://npisforum.eu/ |
| | 14 | Support adherence to the guideline | NPIS Model Glossary<br>https://npisociety.org/glossaire/#<br>NPIS Regitry<br>https://www.referentielinm.org/en/ |

| | | | |
|---|---|---|---|
| | 15 | Evaluate the impact of the reporting guidance | NPI Society |
| | 16 | Develop Web site | NPI Society https://npisociety.org/npismodel |
| | 17 | Translate guideline | NPIS Model is available in English, Spanish, French |
| | 18 | Update guideline | NPI Society + future work |
